# Bridging individual and population dynamics of acute respiratory viruses to optimize antiviral interventions in nursing homes

**DOI:** 10.1101/2025.11.15.25340292

**Authors:** Hind Zaaraoui, Clarisse Schumer, Xavier Duval, Sylvie van der Werf, Gaetan Gavazzi, Nathan Peiffer-Smadja, Lulla Opatowski, Jérémie Guedj

## Abstract

Respiratory virus outbreaks impose a recurrent burden in nursing homes, where elderly residents face high risks of infection and severe complications. Although antiviral treatments exist, their use remains limited due to insufficient evidence on efficacy against virus transmission. In this context, outbreak control often relies on contact precaution and isolating symptomatic residents and/or their contacts.

To evaluate pharmacological interventions, we developed a multiscale model integrating within-host viral kinetics, contact patterns, and transmission dynamics in nursing homes. The framework was applied to outbreaks caused by SARS-CoV-2, influenza virus (IAV), and respiratory syncytial virus (RSV) in nursing home populations. Calibrated to reflect typical nursing home individual interactions and the activity of antivirals reducing residents’ severity by 50%, the model shows that treating symptomatic individuals together with their close contacts could markedly reduce infection burden. In such scenarios, incidence of severe cases and overall cases decreased by up to 80% and 45–70%, respectively. Notably, such strategies were predicted to be at least as effective as isolation- and contact-reduction–based measures. In contrast, treating only symptomatic residents alone had limited impact on transmission, because of the strong role of pre- and asymptomatic transmission, especially for SARS-CoV-2. Antivirals with modest efficacy were unlikely to substantially mitigate SARS-CoV-2 outbreaks but could still prove effective against IAV and RSV, where transmission dynamics are more favorable.

This modeling approach combines within- and between-host processes to provide a comprehensive understanding of respiratory virus outbreaks in nursing homes. It can guide the design of targeted interventions, optimize antiviral deployment, and reduce reliance on isolation, thereby helping to mitigate both outbreak size and severity.

## Introduction

Nursing home residents are among the most vulnerable populations to viral infections due to a combination of factors, including advanced age, weakened immune responses, and a high prevalence of comorbidities Mehta et al. [2021], Rutten et al. [2020], Jeandel and Guérin [2021]. Additionally, the communal living environment and the existence of care staff who interect with many residents create ideal conditions for the rapid spread of infectious diseases, particularly respiratory viruses such as SARS-CoV-2, influenza and respiratory syncytial virus (RSV). These infections pose a significant threat, often leading to severe complications, hospitalizations, and increased mortality rates Landi et al. [2024], Rutten et al. [2020], Utsumi et al. [2010], Gaillat et al. [2008]. Given the fragility of this population and the limited effectiveness of natural immune responses, the need for proactive interventions is critical.

To mitigate the spread of infections in nursing homes, drastic isolation measures have been widely implemented during the first waves of the COVID-19 pandemic, including prolonged lockdowns, strict visitor restrictions, and limited communal activities Dujmovic et al. [2022]. While these measures have effectively reduced viral transmission, they have also had severe consequences on residents’ mental and physical health San Martín-Erice et al. [2024], Simard and Volicer [2020], Dujmovic et al. [2022]. Isolation, reduced physical activity, and lack of family interactions have been associated with cognitive decline, depression, anxiety, and physical deconditioning, further compromising the overall well-being of this already vulnerable population Levere et al. [2021]. This highlights the urgent need for alternative strategies that balance infection control with maintaining residents’ quality of life.

Among the most promising interventions, antiviral agents and monoclonal antibodies (mAbs) have emerged as critical tools for both the prevention and treatment of viral infections in nursing home populations Ma et al. [2023], Cohen et al. [2021], Worcel et al. [2021], Van den Dool et al. [2009], van der Sande et al. [2014]. When administered in a timely manner, these therapies can substantially reduce the risk of progression to severe disease among residents. Although the concept of early treatment to limit transmission is well established, its implementation has historically been hindered by the absence of effective therapeutic options and robust clinical trial evidence. However, this landscape is evolving with the recent development of efficacious antivirals targeting SARS-CoV-2, influenza (IAV), and RSV.

Beyond protecting infected individuals, the use of antivirals and monoclonal antibodies in nursing homes can also contribute in curbing viral transmission Cohen et al. [2021]. Treating both symptomatic residents and their close resident contacts can help reduce viral shedding in infected individuals, limiting the spread of the virus to other residents and nursing home workers (NHW). This approach is particularly crucial in environments where complete isolation of infected individuals is challenging and where vaccination alone may not provide sufficient protection due to declining immunity or emerging viral variants van Ewijk et al. [2022], Pop-Vicas et al. [2015], Peng et al. [2025].

To better understand and quantify the impact of such antiviral-based interventions, this article introduces a multi-scale model that links within-host viral dynamics to between human transmission Zaaraoui et al. [2024] within a nursing home for SARS-CoV-2, influenza and RSV, and the probability of severe disease. By integrating individual-level viral kinetics with population-level infection spread and risk assessment, this model provides a comprehensive framework for evaluating how antiviral and antibody-based treatments influence both disease severity and outbreak control in nursing home settings. We use that approach to help optimize treatment strategies, with the ultimate goal to reduce the burden of severe infections in long-term care facilities, while maintaining at maximum some continuity in social activities/interactions.

## Methods

### Within-host model

#### Viral dynamics

We used previously developed viral kinetic models in the Upper Respiratory Tract (URT) for SARS-CoV-2 Zaaraoui et al. [2024], influenza (IAV), and respiratory syncytial virus (RSV) (see Supplementary). In brief, the model incorporates three cell populations: susceptible target cells (*T*), non-productively infected cells (*I*_1_) and productively infected cells (*I*_2_). The target cells are infected at rate *β*, transition to productively infected at rate *k*, and are cleared at rate *δ*, producing virus at rate *p*. In the case of COVID-19, a fraction *µ* of virions is infectious (*V*_*I*_), while the remainder is non-infectious (*V*_*NI*_), both cleared at a rate *c*, with a total viral load *V* (*t*) = *V*_*I*_(*t*) + *V*_*NI*_(*t*). To simplify the new models for both IAV and RSV, we do not differentiate between infectious and non-infectious viral particles. An immune response compartment *F*, induced by the presence of viral antigens, reduces viral production in a non-linear fashion 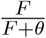 and decays at a rate *d*_*F*_ . For SARS-CoV-2 and RSV, a refractory compartment *R* is also modeled to accounts for cells that cannot be infected (see Supplementary and parameter values in Table S1).

In addition, we incorporated in the model an age-related immune impairment to mimic prolonged viral shedding in the elderly population. This was modeled via a 25% lower loss rate of infected cells, leading to an extended duration of viral shedding of *≈* 4-5 days Zhou et al. [2020], Noh et al. [2014].

In symptomatic individuals, the incubation period was assumed to be log-normally distributed with a median duration of 4, 2 and 5 days for SARS-CoV-2, IAV, and RSV, respectively Zaaraoui et al. [2024], Wu et al. [2022], Kelly et al. [2016].

#### Severe disease

We assumed that 50% of infected individuals remain asymptomatic Rutten et al. [2020]. In the 50% of symptomatic infected individuals, we modeled the risk of severe disease (e.g., need for supplemental oxygen, hospitalization or death) in residents as follows. We used a Weibull model to relate the instantaneous risk of severe disease at time *t* and the viral load at time *t*− 4, *V* (*t*− 4), to reflect the delay between the viral load in the upper and the lower respiratory tract (LRT) Ke et al. [2020]:

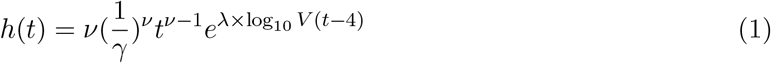

where *λ* quantifies the independent effect of viral load in LRT on severe disease, while *ν* and *γ* are scaling parameters. The cumulated risk of severe disease at time *t, R*(*t*), is then obtained as :

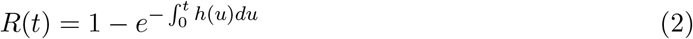

Parameters in Eq.1 were chosen to reproduce typical risk of severe disease in symptomatic individuals for SARS-CoV-2, IAV and RSV, 25%, 12% and 12%, respectively Costemalle et al., Sousa et al. [2020] (see Table S1).

#### Antiviral treatment

We incorporated in the model the initiation of a putative antiviral treatment at time *t* = *t*_*x*_ reducing viral production with an efficacy *ϵ*, and we further assumed that treatment is continued until virus eradication. In our main scenario, *ϵ* was calibrated for each virus to reproduce a 50% reduction in the risk of severe disease when administered shortly after symptom onset (within 5 days for SARS-CoV-2 and RSV, 2 days for IAV). This corresponds to *ϵ* equal to 0.94, 0.996 and 0.84 for SARS-CoV-2, IAV and RSV (see above and Table S1 and S2). Sensitivity analyses were conducted with different values of *ϵ* corresponding to clinical efficacy of 80% and 30%.

### Contact model in the nursing home

#### Frequency and duration of contacts

The frequency and duration of at-risk contacts that can lead to transmission between individuals within the nursing home depend on individuals’ characteristics. We define here two contact matrices that quantify (1) the contacts, ℱ, and (2) the duration of contacts per day, *D*, between individuals of different categories, residents and nursing home workers (NHW).

The matrices were parameterized based on empirical matrices Duval et al. [2018] built from data collected in two geriatric ward of a Long Term Care Facility (LTCF) in France, where wearable sensors were distributed to all individuals to collect close proximity interactions of less than 1.50m.

Focusing on two geriatric wards, we used the estimates of average total duration of contact (in minutes) and average number of contacts per day (number of individuals with whom one comes into contact per day) between residents and NHW.

The average group-level (residents and NHW) number of distinct contacts per day *F* between NHW and residents was:

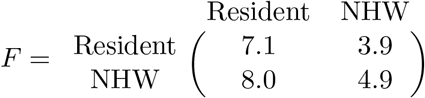

The corresponding group-level matrix of mean cumulative daily contact durations *D* (in minutes) is given by:

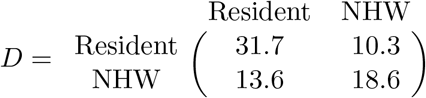

Based on these matrices, a *N ×N* general contact matrix *C*(*t*) was stochastically generated each day. This matrix modeled, for each *i* and *j*, the cumulative duration of contact between individuals *i* and *j* on that day. For each (*i, j*) and *t, C*_*i,j*_(*t*) is generated to match, on average, the empirical contact frequencies and average duration of contacts by categories. Details on the construction of *C*(*t*) is provided in Supplementary Methods.

Moreover, previous studies reported a large proportion of recurrent contacts in these settings, meaning that individuals in contact on one day had more chance to meet again on the next day Duval [2019], Vanhems et al. [2013]. This feature was accounted for in the calculation of *C*(*t*), through the definition of a time dependent weight *ω*_*i,j*_(*t*) that advantaged contacts with individuals already encountered the day before and the previous days (see details in Supplementary Methods).

#### Risk of transmission during a contact

As in Zaaraoui et al. [2024], Ke et al. [2021], we used a Power-law to model the (non-linear) relationship between viral load in individual *i* and his/her risk of transmission to any other individual over a 5-minute contact. We defined *p*_*i*_(*t*) by:

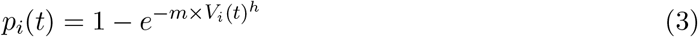

where *m* quantifies the strength of the association between viral load and transmission, and *h* reflects the stiffness of this association, fixed to *h* = 0.49 Ke et al. [2021], Zaaraoui et al. [2024].

To compute the effective transmission risk between from *i* to any individual *j* on day *t*, the contact duration per day *C*_*i,j*_(*t*) was discretized by intervals of 5 minutes. Contact durations of less than 5 minutes were associated to a null probability of transmission. On the other side, transmission risk was assumed to saturate after 30 minutes of contact per day. The effective probability that an individual *i* transmits the infection to individual *j* during contacts occurring on a given day *t* is obtained by integrating the instantaneous transmission probability over the cumulative daily duration of contact between individuals *i* and *j* on that day, given by the time-dependent contact matrix *C*_*i,j*_(*t*) (see Supplementary Methods). Accordingly, the transmission probability on day *t* given a contact between *i*, infected, and *j*, susceptible, is expressed as:

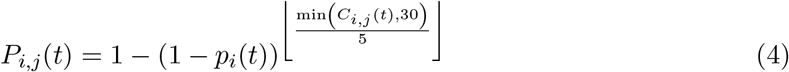

where *p*_*i*_(*t*) is the instantaneous transmission probability at day *t* defined in (3) for the infected individual *i*. In the simulations, infection of *j* was randomly sampled into a Bernoulli distribution of parameter *P*_*i,j*_(*t*).

#### Link with the basic reproductive number

The nursing home reproductive number, noted *R*_0_, was calculated by simulating 1,000 outbreaks and computing the average number of secondary cases (residents and NHW) from an infected index case in the nursing home. The transmission model described above was calibrated in order to match realistic *R*_0_ values for the investigated viruses. The value of parameter *m* was adjusted, after integration of the contact patterns, to reach on average *R*_0_ = 3, 1.5 and 1.5 for SARS-CoV-2, IAV and RSV, respectively Liu et al. [2020a,b], Biggerstaff et al. [2014] (see Table S1).

### Simulation of interventions

#### Nursing home characteristics

All scenarios were assessed by simulating the dynamics of viral infections following one introduction in 1000 independent nursing homes of size *N* = 120, and average NHW/resident ratio of 0.73, consistent with French statistics DREES [2023].

#### Intervention strategies and evaluation of their effectiveness

Following the introduction of an index case, simulations reaching two or more symptomatic residents, i.e., at least one transmission event in residents were defined as “outbreaks”. By opposition, simulations where no secondary transmission occurred in residents were considered as “extinctions”. In all scenarios, interventions were implemented within 5 days from symptom onset of the secondary symptomatic case. The following interventions were assessed, alone or in combination:

- **Mask facing in NHWs with surgical masks**, assumed to reduce by 80% viral transmission (in masked transmitter) and viral infection (in masked susceptible) Banholzer et al. [2023], Liang et al. [2020].
- **Isolation of residents**, assumed to eliminate all between-resident contacts and reduce residents-NHWs contacts by 80%. Three distinct isolation strategies were considered: isolating symptomatic residents only for 10 days; isolating symptomatic and their contacts for 10 days; or isolating all nursing home residents until outbreak extinction (isolation ends 10 days after detection of the last symptomatic).
- **Treatment of symptomatic residents**. In this scenario, antiviral treatment is administered within 5 days from symptom onset of any symptomatic resident. It is assumed to be continued until viral clearance.
- **Treatment of positive contacts (residents)**. This scenario assumed that after detection of the index case, contact residents are tested. Antiviral treatment is then administered to the index case and to his/her contacts in case of positive virological test (viral load greater than 2.7 log_10_ for SARS-CoV-2, and 0.7 log_10_ for IAV and RSV). Similarly, treatment is assumed to continue until viral clearance.
- **Treatment of all contacts (residents)** Antiviral treatment are administered to the index case and all his/her contacts regardless of their infection status. Two different drugs were considered: short half-life oral antiviral drugs or monoclonal antibodies with long half-life. In case of monoclonal antibodies, constant antiviral activity is assumed from the time of administration to the end of the outbreak. In case of oral antiviral drug, drugs were active for 5 days only in contacts, and treatment can be administered several times in case of repeated exposures. Note that in this setting, we also assumed that uninfected individuals were fully protected against infection acquisition for 5 days.

The listed intervention strategies were compared based on the following endpoints, after the last case of the outbreak:

- Infection attack rate, defined by the proportion of residents infected.
- Severe infection attack rate, defined by the proportion of residents with a severe infection.
- Isolation duration, defined by the average number of isolation days in residents.
- Treatment intake, defined by the average number of treatment courses in residents.

## Results

### Quantifying the role of viral dynamics on infection and transmission

The within-host model (see Supplementary Methods) reproduces the large variability of viral kinetics in the general population during SARS-CoV-2, IAV and RSV infection, with a median time to peak viral load of 4, 2 and 6 days, respectively, and a median time to clearance of 17, 6 and 12 days after infection, respectively. The model incorporates a less efficient immune control in elderly, leading to a time to viral clearance of 22, 10 and 17 days, respectively, in the population of residents (Fig. 1). The model assumes that symptom and viral kinetics occur independently, and that 50% of infected individuals are symptomatic. In symptomatic individuals, the amount of virus shed during the pre-symptomatic phase of the infection differs greatly between viruses, with the AUC of virus shed before symptom onset representing 40%, 60% and 30% of the total amount of virus shedding for SARS-CoV-2, IAV and RSV respectively.

**Figure 1.**
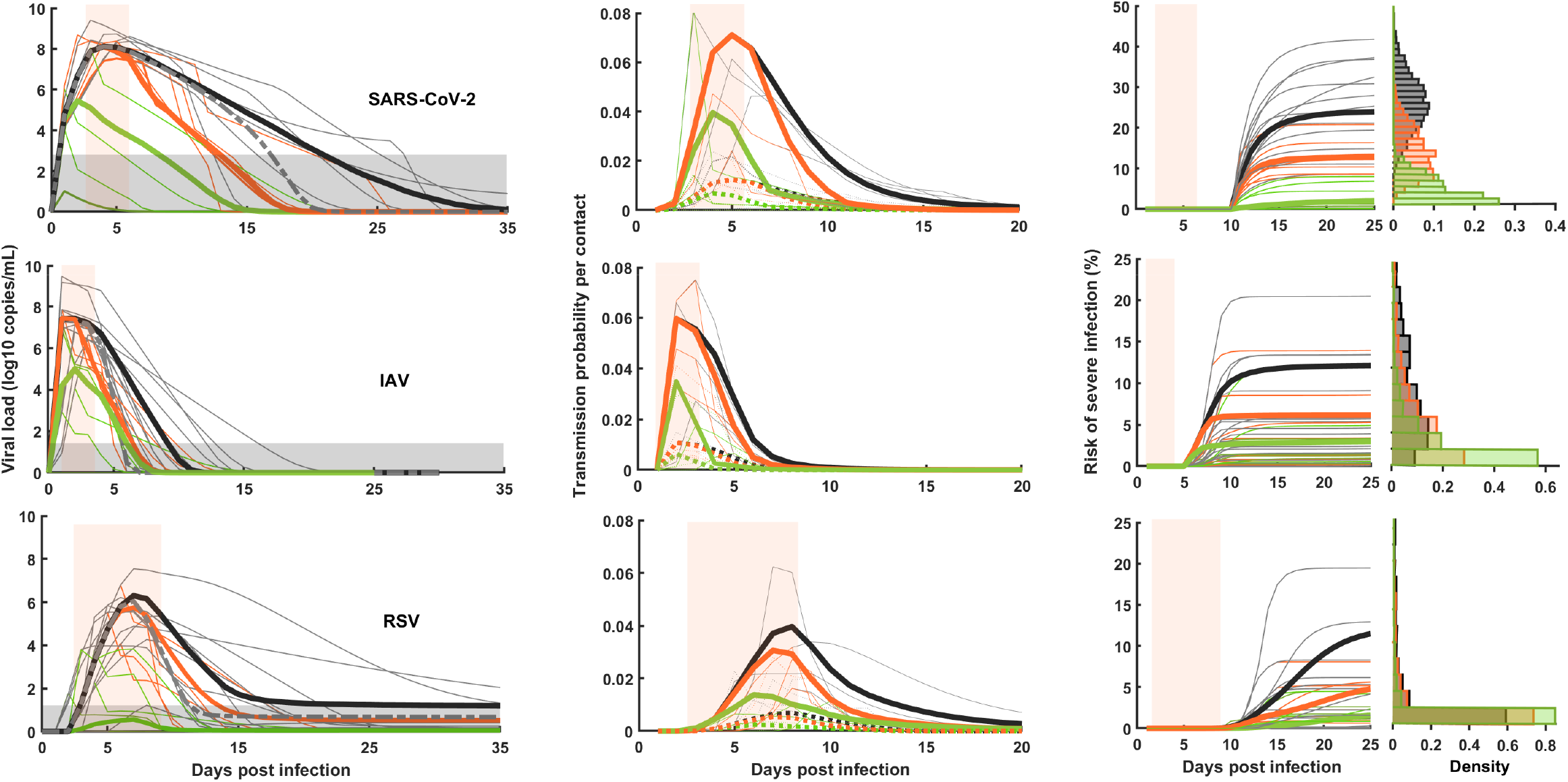
Viral load, transmission probability dynamics and risk of severe disease in residents. **Left column** Viral load kinetics (NHW median viral load in dashed line); **Center column** Probability of transmission over a punctual contact (5 min) in dashed lines, and 30 minutes per contact in continuous lines; **Right column** Risk of severe infection. Black: no treatment; orange: treatment after symptom onset; green: treatment before symptom onset. The light curves represent typical evolution without treatment, and the bold are median kinetics. Gray bold dashed curves in left figures represent viral load dynamic for NHW. Orange shaded area: incubation period.

The model also captures the importance of the timing of treatment initiation, with post-exposure prophylaxis being more effective than treatment administered after peak viral load (Fig. 1). The clinical model relates the risk of severe infection (i.e., hospitalization) to the viral dynamics using a time-to-event model ((2)). Treatment efficacy was calibrated to reduce the risk of severe infection by 50% when administered shortly after symptom onset (see Methods). Administration of treatment before symptom onset further increases the efficacy, although with a different magnitude for the three viruses (Table S2). The transmission model relates the risk of virus transmission during a contact at time *t* to the viral load using a probabilistic model (3). Because the risk of transmission also depends on the contact patterns, a model of contact was developed to reproduce the typical structure of contact in a typical nursing home of 120 people (68 residents and 52 NHW) (see Fig. 2 and Table S2). In that perspective, administration of treatment also reduced the instantaneous risk of transmission during high-risk contact, from about 20-30% when administered after symptom onset to 60-80 % when administered before symptom onset (Table S2).

**Figure 2.**
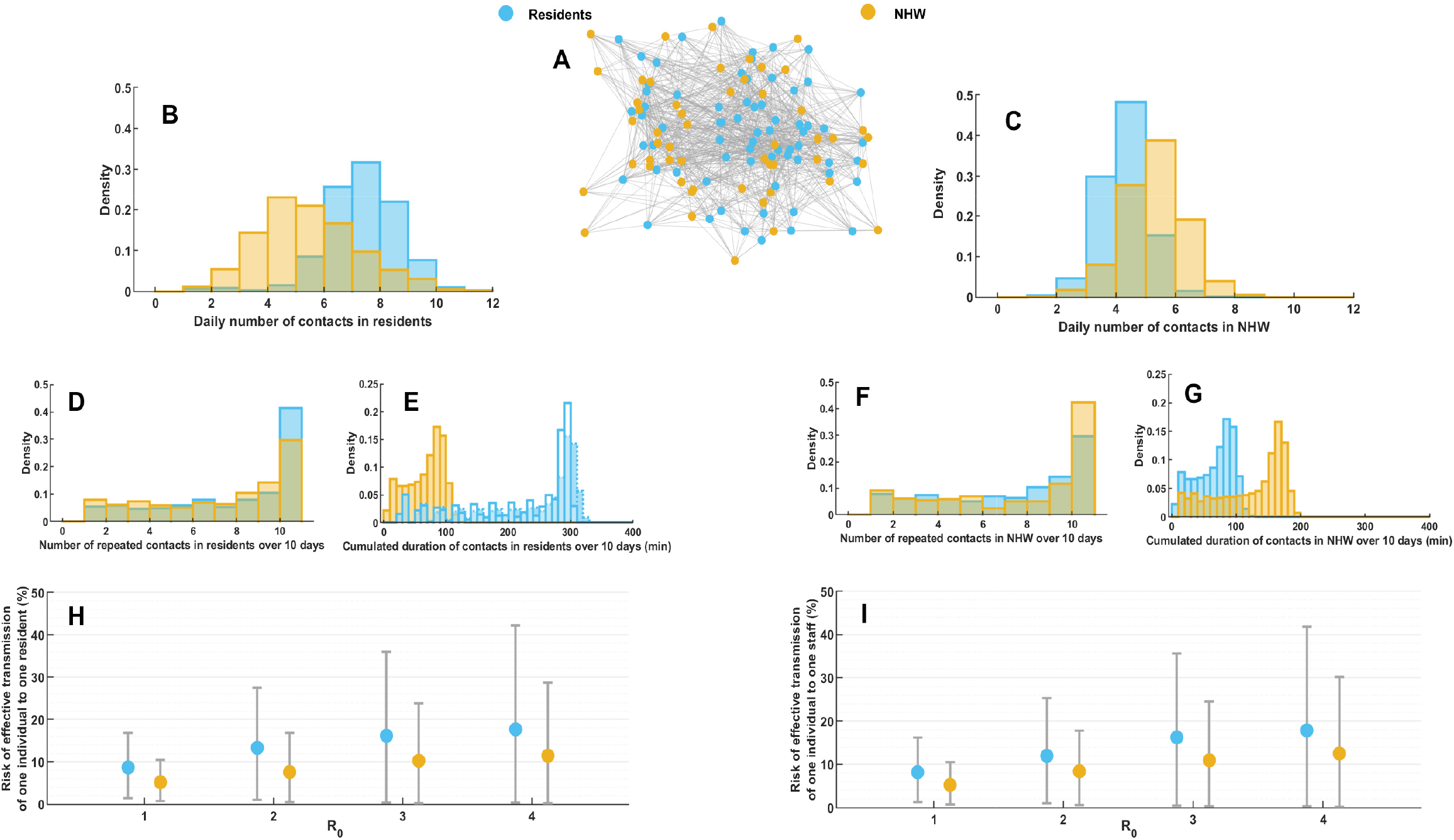
Contacts patterns between residents and NHW. **A**: Typical network of contact in the nursing home in a day. **B**: Number of individuals in contact with a resident in a day. **C**: Number of recurrent contacts for a resident over 10 days. **D**: Cumulated duration of the contacts for a resident over 10 days; A transparent distribution of these contact durations is added to represent the saturated durations up to 30 min which are taken into consideration in our simulations. **E**: Risk of transmission by a resident to any other individual. **F-I** Same as B-E in NHW.

### Comparison of different strategies to reduce outbreak size

#### Non-pharmacological interventions

For each strategy and each virus, we simulated a large number of independent simulations in nursing homes. Nursing homes with outbreaks represented 90%, 40% and 35% of simulations for SARS-CoV-2, IAV and RSV, respectively. In absence of any interventions, the size and the impact of the outbreak are directly related to the risk of transmission and the risk of severe disease, respectively. Using the parameter values of the model, the mean infection attack rate was equal to 97, 58 and 59 % for SARS-CoV-2, IAV and RSV, respectively, which, consistent with our assumptions on disease severity, led to a mean severe infection attack rate of 12.5, 3.6 and 3.6% (see Fig. 3).

**Figure 3.**
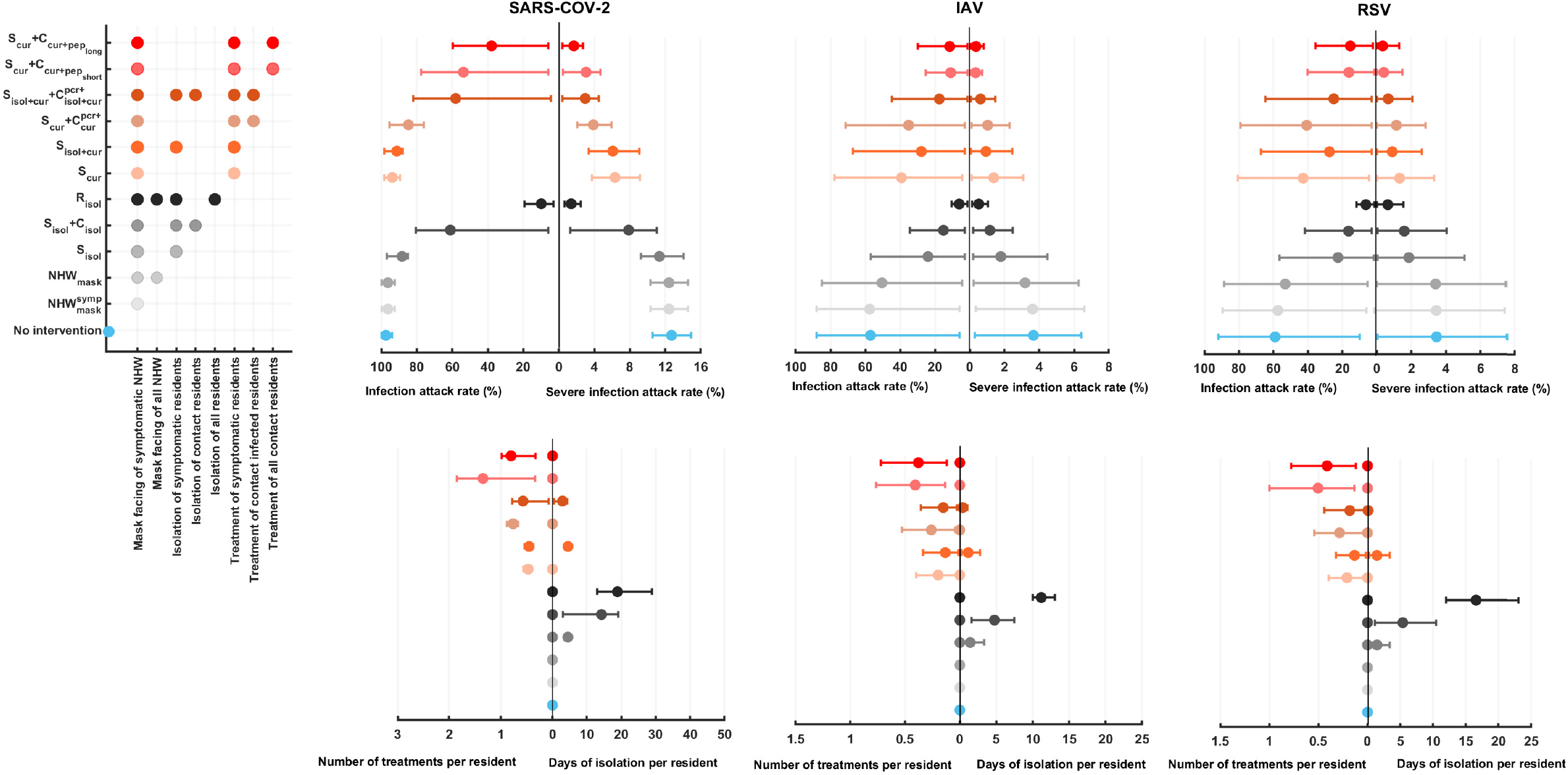
Impact of intervention strategies on outbreak severity with the number of treatments and of isolation days per residents needed. **Up**: Percentage of infected residents and severe infection cases. **Bottom**: Number of isolation days and number of treatments per resident in nursing homes. Circles represent the average of the infection and severe infection attack rates, number of isolation days and treatments per residents (IQ: 10%-90%).

The most obvious strategies to contain an outbreak rely on non-pharmacological measures, such as isolation of residents or mask facing. Mask facing was assumed to reduce both the risk of transmission and infection by 80% (see Methods). In the rest of the manuscript, and unless stated otherwise, mask facing of symptomatic NHW, noted 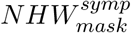, was considered as the baseline reference intervention, and led to a mean infection attack rate equal to 95, 57 and 57% for SARS-CoV-2, IAV and RSV, respectively, while the mean severe infection attack rate was 12.4, 3.5 and 3.5%, i.e., very close to results obtained without any intervention. Universal masking of NHWs upon outbreak, irrespective of symptoms, *NHW*_*mask*_, led to similar results.

Isolation of symptomatic cases only,*S*_*isol*_, had only limited effectiveness (Fig. 3). In fact *S*_*isol*_ had a nearly similar effectiveness than *NHW*_*mask*_, suggesting that in our framework non-symptomatic NHW and symptomatic residents had similar contribution to overall transmission. In order to effectively stop transmission, isolation-based strategies need to focus on a- and pre-symptomatic residents. Indeed isolation of all residents, regardless of symptoms, and universal mask facing, *R*_*isol*_, reduced the infection and severe infection attack rates to 10% and 1.5% respectively for SARS-CoV-2, and 5% for IAV and RSV, i.e., an effectiveness of more than 85% for the three viruses.

#### Treatment of symptomatic residents

For the same reason, strategies relying on treatment of symptomatic residents, *S*_*cur*_ and *S*_*isol*+*cur*_ has modest effectiveness on transmission (see Fig. 4), in particular for SARS-CoV-2, where most transmissions occurred before the initiation of treatment in these individuals. The effectiveness was larger for IAV and RSV, characterized by a lower contribution of pre-symptomatic transmission, with an effectiveness estimated to about 20%. The effectiveness was however larger when looking at the outbreak severity, as treatment reduces by 50% the risk of severe disease in symptomatic individuals. Overall, *S*_*cur*_ reduced the severe infection attack rate with an effectiveness of 50, 62 and 60% after SARS-CoV-2, IAV and RSV outbreaks, respectively (Fig. 4).

**Figure 4.**
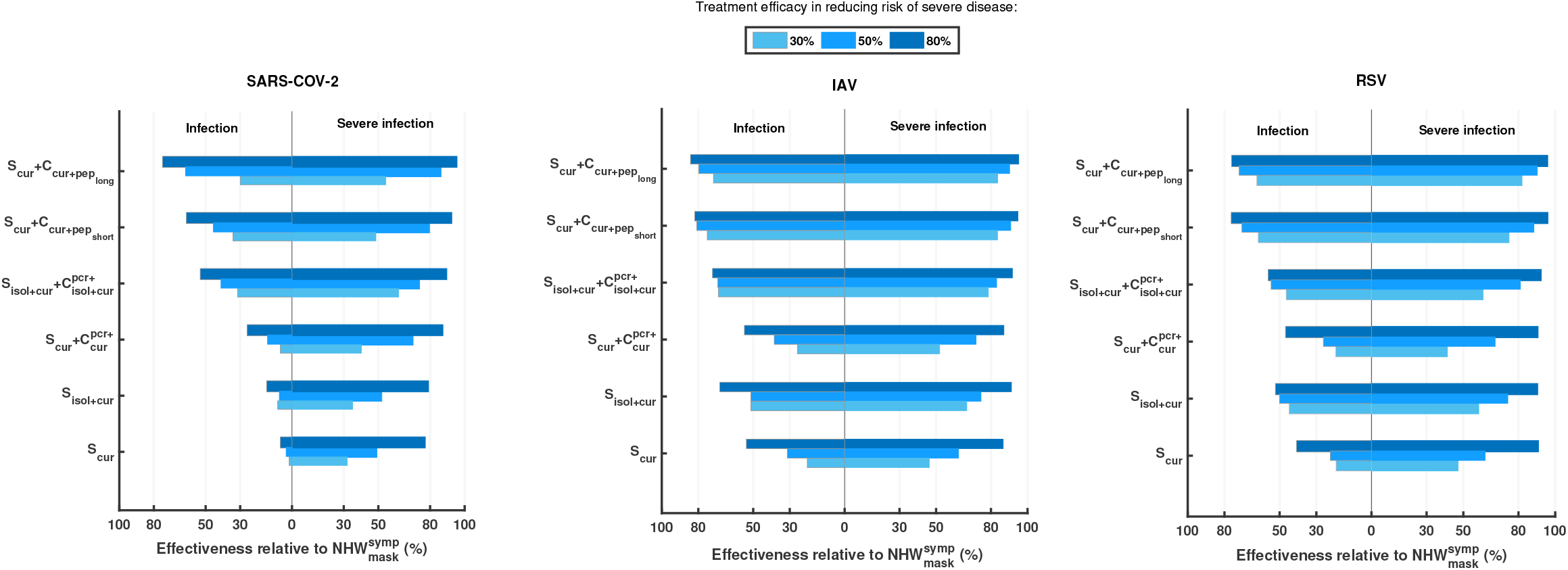
Relative efficacy of intervention strategies on the risk of hospitalization for different values of antiviral treatment efficacy, and respective mean isolation days and number of treatments per resident. The reference strategy is mask wearing for symptomatic NHW 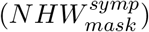.

#### Interventions based on symptomatic and contact residents treatment

Next, we considered strategies focusing on treating not only symptomatic individuals, but also their resident contacts.

##### Treatment of the index and positive contacts

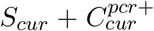, effectively reduced transmission of IAV, with an effectiveness of 39% (see Fig. 4), but was much less effective against SARS-CoV-2 and RSV (less than 25%). This is due to the fact that the time between virus exposure and PCR positivity is only 1 day with IAV and about two days for RSV and SARS-CoV-2, which increases the risk of false negative results. Accordingly, for these two viruses, there was a benefit of also isolating PCR positive individuals, with an effectiveness on transmission of 40 and 55%, respectively. These two strategies were highly effective in reducing the number of severe cases, with effectiveness larger than 74, 83 and 81% for SARS-CoV-2, IAV and RSV, respectively.

##### PEP treatment strategies

Finally, we examined a treatment strategy in which all contact residents receive post-exposure prophylaxis (PEP) or curative treatments, regardless of their infection status as determined by virological testing. As treatment may either rely on oral antiviral drugs with short half-life (e.g., Nirmatrelvir + Ritonavir, Oseltamivir, Baloxavir marboxil), or injected monoclonal antibodies with longer half-life (e.g., Tixagévimab + Cilgavimab, Nirservimab), we considered treatment protecting only for 5 days, noted 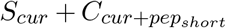, but also treatments protecting for several weeks, 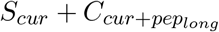. The effectiveness obtained with long-half life drugs were large, with protection against transmission equal to 62, 80 and 72% for SARS-CoV-2, IAV and RSV, respectively. The differences with oral antiviral drugs were small for IAV and RSV, with effectiveness on transmission equal 70 and 68% respectively, and a lower effectiveness of 45% for SARS-CoV-2, where 5 days protection is not sufficient due to a slower viral clearance. Both strategies led to high protection against severe cases, with an effectiveness larger than 80% for all viruses. These results are close to those obtained with full isolation, showing that such strategies represent real alternative to isolation-based strategies.

### Implications of these strategies on outbreak duration, isolation of residents and number of treatment courses

#### Duration of outbreak and isolation

We finally examined the effect of these different strategies on the typical duration of an outbreak, defined as the time from its detection to the last symptomatic case in residents. The reference intervention, 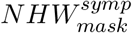, led to a mean duration of 5, 3.5 and 3.5 weeks for SARS-CoV-2, IAV and RSV, respectively, close to that obtained in absence of any intervention. Effective strategies relying on isolation or treatment of contacts shortened outbreak duration to 2 weeks for IAV and RSV, with no considerable difference for SARS-CoV-2. The strictest isolation strategy considered, *R*_*isol*_, could not fully prevent the transmission of highly transmissible virus occurring via NHW, and the mean duration of outbreak was equal to 3 weeks for SARS-CoV-2. Accordingly, this implies that strategies relying on isolation of symptomatic or contact cases need to be implemented for long period of times to successfully contain an outbreak. For instance, *S*_*isol*_ + *C*_*isol*_ was associated with a mean duration of isolation per resident of 14, 4.8 and 5.4 days for SARS-CoV-2, IAV and RSV, respectively, and reaching up to 19, 11 and 17 days for *R*_*isol*_ (Fig. 4). Yet, *S*_*isol*_ + *C*_*isol*_ effectiveness in terms of number of infected cases was not larger than that based on treatment without isolation such as 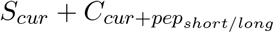 (see Fig. 3).

#### Number of individuals treated

Obviously, the benefit of treatment based strategies need to be put in perspective with the efforts in terms of treatment deployment. This is particularly true when viruses are highly transmissible for longer periods, such as SARS-CoV-2, and that treatment have a short half-life. Strategy 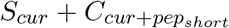 had a mean number of treatment per resident equal to 1.3, 0.4 and 0.5 for SARS-CoV-2, IAV and RSV, respectively (see Fig. 4). This number could be substantially reduced by using treatment conferring long protection, such as monoclonal antibodies. In this case this average number of treatment was reduced to 0.8, 0.35 and 0.4 for SARS-CoV-2, IAV and RSV, respectively.

### Sensitivity analysis - exploring different levels of antiviral efficacy

In a last step, we assessed the benefit of treatment based strategies for different levels of antiviral efficacy, considering lower clinical efficacy (30%, low efficacy) or higher efficacy (80%, high efficacy). Interestingly, for viruses with relatively low transmission rates (such as IAV or RSV), a strong reduction in transmission amplified the clinical effect of treatment on the risk of severe disease, resulting in an overall synergistic impact exceeding the clinical efficacy of the treatment. These findings underscore the advantages of such combined approaches at both the individual and population levels. This was particularly true in the case of less transmissible viruses (IAV or RSV) with efficacy against severe disease attack rate above 50% in almost all scenarios considered (including low treatment efficacy), and that could reach more than 90% in cases of highly effective treatment Fig. 4). In general, the effects of treatment against transmission were relatively insensitive to antiviral treatment efficacy, in particular when the contacts were treated. In this case, the effectiveness against transmissions increased with the level of antiviral activity, reaching values as large as 75% or higher for all three viruses. Considering the effectiveness in reducing the severe infections, since the effectiveness of treatments on transmission were high, the values exceed the clinical efficacy of the treatments particularly for IAV and RSV. Treatments with a clinical efficacy of 30% achieve more than a 50% reduction in the risk of severe infections across all strategies, and reach approximately 80% in strategies that include treatment of contacts. Except for SARS-CoV-2, varying the clinical efficacy of treatments has little influence on overall effectiveness for any strategy, including those involving contact treatment.

In terms of intervention burden, isolation days were minimally affected, particularly when comparing 30%–50% clinical efficacy (see Fig. S1). For COVID-19, the number of treatments administered per resident exhibited greater variability under PEP strategies, with relative changes of 20% and +10% at 80% and 30% treatment efficacy in reducing severe disease, respectively, compared with the 50% efficacy reference scenario. More results for IAV and RSV and for all the strategies are presented in Fig. S1.

## Discussion

We here developed a multiscale model integrating within-host viral kinetics and between-host transmission dynamics to simulate respiratory infection outbreaks in nursing homes. By combining empirical contact data Duval et al. [2018], Duval [2019] with the mechanistic models of infection, our framework captures the complexity of outbreak dynamics and enabled us to assess individual and population effects of different strategies of outbreak mitigation. The model was calibrated to reproduce typical viral kinetics and transmission observed with SARS-CoV-2, IAV and RSV. The three viruses differ in their viral kinetic patterns (time to peak, shedding duration) but also their infectiousness (with SARS-CoV-2 being much more transmissible). These differences enabled us to tease out the different factors that may modulate the outbreak. In our simulations, the duration of outbreaks, defined as the period until recovery of the last case, whether staff or resident, was estimated at approximately five weeks for SARS-CoV-2, and three weeks for both IAV and RSV. These findings are consistent with previously reported outbreak durations in long-term care facilities, where COVID-19 outbreaks were often prolonged, with median durations ranging from three to six weeks depending on setting and control measures Office [2022], while influenza and RSV outbreaks generally resolved within two to three weeks Ferrante et al. [2024], Yip et al. [2018], Millership and Cummins [2015]. The simulated attack rates, which included both symptomatic and asymptomatic infections, were more than 80% for COVID-19 and 50% for both IAV and RSV. These estimates align with published observations of high COVID-19 attack rates in nursing homes during uncontrolled outbreaks, up to 87% Reeve et al. [2023], and attack rates for influenza and RSV typically ranging from 30% to 60% in such settings Lansbury et al. [2017], Osei-Yeboah et al. [2024]. Taken together, these results support the validity of our modeling framework, reflecting outbreak dynamics observed in real-world long-term care facilities, and highlighting both the severity of COVID-19 in these environments and the substantial, though generally shorter, impact of influenza and RSV outbreaks.

Our model emphasized the synergetic effects of antiviral treatment to both reduce the risk of severe disease and the number of infected individuals. Interventions that included pharmacological treatment of symptomatic individuals and their contacts consistently achieved large reduction in both the number of infected individuals, ranging from 45 to 70% (depending on the virus transmissibility) and about 80% in the number of severe cases. In contrast strategies focusing only on symptomatic cases had very little effect on transmission, as symptom occur close or after peak viral load. This result is in line with another modeling work that did not include within-host viral kinetics, showing that antiviral prophylaxis for influenza in nursing homes for both cases and contacts can reduce morbidity and hospital burden more effectively than case-only strategies Morris et al. [2024]. Interestingly such strong effects on both transmission and risk of severe disease were also found in case of less potent antiviral drug (associated with a clinical efficacy of 30%, instead of 50% in our main scenario), at least for IAV and RSV, but not for SARS-CoV-2. This is due to the fact the effectiveness of antiviral treatment on transmission depends on the force of transmission, which was larger for SARS-CoV-2 (R0=3) than for IAV and RSV (R0=1.5). Further, the effectiveness of these strategies was systematically comparable or even better to isolation-based strategies, supporting the use of antiviral-based strategies as an alternative to isolation.

While the model emphasizes the synergetic effects of antiviral treatment when they are administered in a timely manner to both symptomatic residents and their contacts, it is important to note that the hypothesis made on their efficacy are not out-of-reach. A statewide cohort study Chambers et al. [2023] reported that administration of monoclonal antibodies (mAbs) to residents with SARS-CoV-2 infection during the Alpha and Delta waves reduced the combined risk of hospitalization and death from 25.3% in untreated individuals to 8.8% in treated individuals, corresponding to a relative risk reduction of 65%. This aligns closely with the 70% effectiveness observed here in strategies involving treatment of contact cases. Molnupiravir and nirmatrelvir/ritonavir, two oral treatments, were administered in nursing home patients during the Omicron BA.2 wave, and showed a reduced risk of severe disease of about 50 % Ma et al. [2023]. Interestingly, in within-host analyses, nirmatrelvir and molnupiravir were found to block viral replication by about 98% Phan et al. [2025], which is close to the value used in our model (94%, see Table S1). This supports the use of a within-host model to capture the effects of antiviral treatment not only on viral load, but also on the risk of transmission. For IAV, we have an antiviral treatment efficacy of about 99.6% that reduces the risk of severe disease by 50%, provided that it is administered with 2 days of symptom onset. This level is same to that observed with oseltamivir or baloxavir Kamal et al. [2015], Hanula et al. [2024]. Accordingly, our model predicts that such treatments could also be effective in reducing transmission, in particular if they are given before peak viral load and symptom onset. We estimated that such treatment could reduce the risk of transmission during a high risk contact by 60 and 30%, if given before or after symptom onset, respectively. This is consistent with the high efficacy found recently in two phases 3 study evaluating baloxavir in household studies, which showed that baloxavir reduces by 90% and 30% the risk of infection and transmission, respectively Monto et al. [2025]. In that context, drugs with long half-life could be also be beneficial, increasing the duration of protection. While logistical constraints may influence the choice between short-acting orally available antiviral drugs and long-acting injectable monoclonal antibodies (mAbs), our analysis indicates that both treatment may have comparable effectiveness against IAV and RSV, with mAbs offering a potential advantage for SARS-CoV-2 due to its extended infectious period.

The model makes a number of hypotheses. First we assumed that nasopharyngeal viral load was the only factor associated with transmission. However, studies reported that other independent individual or environmental factors may impact transmission, such as the type of symptoms, or fomite transmission Kraay et al. [2018]. Further the model integrated patient variability in transmission rate, but did not specifically integrate vaccination, which plays a central role in prevention strategies, in particular for SARS-CoV-2 and IAV. While the effect of vaccination on the risk of severe disease is often well known, its effect on viral load, and hence on susceptibility to infection and transmission during a breakthrough infection is poorly known. The role of other factors than viral shedding could also play a role to investigate other strategies, such as ventilation, which have shown promising results to reduce infection Organization [2021]. The contact network data used in our model was collected from real long term care facility observations Duval et al. [2018], which may be different from what is observed in nursing homes in absence of outbreak. Nursing homes in addition are highly diverse, which interactions that can depend on the demographics of the residents, their cultural background, or the number of NHW. Real-world implementation of contact-based treatments may pose operational challenges, including timely identification of contacts, logistical distribution of therapeutics, and resident consent — all of which must be addressed to translate modeling insights into effective public health action. Our model does not integrate the fact that several virus introductions may occur every season, which poses the question of the safety and the feasibility to administer several treatments within a short period of time. Additionally, it does not account for potential drug–drug interactions, particularly relevant for treatments such as Nirmatrelvir combined with Ritonavir, which could limit its use in highly poly-medicated populations. Furthermore, the model does not consider the possible occurrence of adverse effects, which could reduce treatment adherence and thus impact overall effectiveness.

To conclude, we here developed a modeling framework that characterizes within-host and between-host dynamics of respiratory viruses during an outbreak in nursing homes. The model can be used to compare different strategies of outbreak mitigation. It shows that treatment of both symptomatic residents and their contact residents could achieve large effectiveness, even with treatment that only reduces the risk of severe disease by 30%. Our findings highlight the limitations of symptom-based treatment and/or isolation in nursing home settings and support a shift toward targeted pharmacological interventions to reduce the spread and severity of respiratory viral infections and preserve the psychosocial integrity of nursing home life.

## Data Availability

All the data has been simulated by the authors

## References

[1] Nicolas Banholzer, Kathrin Zürcher, Philipp Jent, Pascal Bittel, Lavinia Furrer, Matthias Egger, Tina Hascher, and Lukas Fenner. Sars-cov-2 transmission with and without mask wearing or air cleaners in schools in switzerland: A modeling study of epidemiological, environmental, and molecular data. PLoS Medicine, 20(5):e1004226, 2023.

[2] Matthew Biggerstaff, Simon Cauchemez, Carrie Reed, Manoj Gambhir, and Lyn Finelli. Estimates of the reproduction number for seasonal, pandemic, and zoonotic influenza: a systematic review of the literature. BMC infectious diseases, 14(1):1–20, 2014.

[3] Laura C Chambers, Huong T Chu, Nickolas Lewis, Gauri Kamat, Taylor Fortnam, Philip A Chan, Leanne Lasher, and Joseph W Hogan. Effectiveness of monoclonal antibody therapy for preventing covid-19 hospitalization and mortality in a statewide population. Rhode Island medical journal (2013), 106(5):42, 2023.

[4] Myron S Cohen, Ajay Nirula, Mark J Mulligan, Richard M Novak, Mary Marovich, Catherine Yen, Alexander Stemer, Stockton M Mayer, David Wohl, Blair Brengle, et al. Effect of bamlanivimab vs placebo on incidence of covid-19 among residents and staff of skilled nursing and assisted living facilities: a randomized clinical trial. Jama, 326(1):46–55, 2021.

[5] Vianney Costemalle, Noémie Courtejoie, Albane Miron de l’Espinay, Équipe EpiCov, Josiane Warszawski, Alexandra Rouquette Meyer, Florence Jusot, Ariane Pailhé, Alexis Spire, Claude Martin, et al. Ála veille du deuxiéme confinement, le systéme de dépistage détectait plus de la moitié des personnes infectées par la covid-19.

[6] DREES. L’enquête auprés des établissements d’hébergement pour personnes âgées (ehpa). https://data.drees.solidarites-sante.gouv.fr/explore/dataset/587_l-enquete-aupres-des-etablissements-d-hebergement-pour-personnes-agees-ehpa/information/, 2023.

[7] Morgane Dujmovic, Thomas Roederer, Severine Frison, Carla Melki, Thomas Lauvin, and Emmanuel Grellety. Covid-19 in french nursing homes during the second pandemic wave: a mixed-methods cross-sectional study. BMJ open, 12(9):e060276, 2022.

[8] Audrey Duval. Comprendre et contrôler la transmission des bactéries multirésistantes par l’analyse et la modélisation des réseaux d’interactions interindividuelles en milieu hospitalier. PhD thesis, Université Paris Saclay (COmUE), 2019.

[9] Audrey Duval, Thomas Obadia, Lucie Martinet, Pierre-Yves Boëlle, Eric Fleury, Didier Guillemot, Lulla Opatowski, and Laura Temime. Measuring dynamic social contacts in a rehabilitation hospital: effect of wards, patient and staff characteristics. Scientific reports, 8(1):1686, 2018.

[10] Christina Ferrante, Christina Bancej, and Nicole Atchessi. Disease burden attributable to respiratory syncytial virus outbreaks in long-term care. Canada Communicable Disease Report, 50(1-2):25, 2024.

[11] J Gaillat, G Dennetiére, E Raffin-Bru, M Valette, and MC Blanc. Summer influenza outbreak in a home for the elderly: application of preventive measures. Journal of Hospital Infection, 70(3):272–277, 2008.

[12] Ryan Hanula, Emilie Bortolussi-Courval, Arielle Mendel, Brian J Ward, Todd C Lee, and Emily G McDonald. Evaluation of oseltamivir used to prevent hospitalization in outpatients with influenza: a systematic review and meta-analysis. JAMA Internal Medicine, 184(1): 18–27, 2024.

[13] C Jeandel and O Guérin. Unités de soins de longue durée et ehpad. Rapport de mission, 25, 2021.

[14] Mohamed A Kamal, Ronald Gieschke, Annabelle Lemenuel-Diot, Catherine AA Beauchemin, Patrick F Smith, and Craig R Rayner. A drug-disease model describing the effect of oseltamivir neuraminidase inhibition on influenza virus progression. Antimicrobial agents and chemotherapy, 59(9):5388–5395, 2015.

[15] Ruian Ke, Carolin Zitzmann, Ruy M Ribeiro, and Alan S Perelson. Kinetics of sars-cov2 infection in the human upper and lower respiratory tracts and their relationship with infectiousness. MedRxiv, pages 2020–09, 2020.

[16] Ruian Ke, Carolin Zitzmann, David D Ho, Ruy M Ribeiro, and Alan S Perelson. In vivo kinetics of SARS-CoV-2 infection and its relationship with a person’s infectiousness. Proceedings of the National Academy of Sciences, 118(49):e2111477118, 2021.

[17] Sean G Kelly, Kristen Metzger, Maureen K Bolon, Christina Silkaitis, Mary Mielnicki, Jane Cullen, Melissa Rooney, Timothy Blanke, AlaaEddin Tahboub, Gary A Noskin, et al. Respiratory syncytial virus outbreak on an adult stem cell transplant unit. American Journal of Infection Control, 44(9):1022–1026, 2016.

[18] Alicia NM Kraay, Michael AL Hayashi, Nancy Hernandez-Ceron, Ian H Spicknall, Marisa C Eisenberg, Rafael Meza, and Joseph NS Eisenberg. Fomite-mediated transmission as a sufficient pathway: a comparative analysis across three viral pathogens. BMC infectious diseases, 18(1):540, 2018.

[19] Suzanne N Landi, Diana C Garofalo, Maya Reimbaeva, Amie M Scott, Lili Jiang, Katherine Cappell, David Lewandowski, Machaon Bonafede, Kaylen Brzozowski, Zuzanna Drebert, et al. Hospitalization following outpatient diagnosis of respiratory syncytial virus in adults. JAMA Network Open, 7(11):e2446010–e2446010, 2024.

[20] Louise E Lansbury, Caroline S Brown, and Jonathan S Nguyen-Van-Tam. Influenza in long-term care facilities. Influenza and other respiratory viruses, 11(5):356–366, 2017.

[21] Michael Levere, Patricia Rowan, and Andrea Wysocki. The adverse effects of the covid-19 pandemic on nursing home resident well-being. Journal of the American Medical Directors Association, 22(5):948–954, 2021.

[22] Mingming Liang, Liang Gao, Ce Cheng, Qin Zhou, John Patrick Uy, Kurt Heiner, and Chenyu Sun. Efficacy of face mask in preventing respiratory virus transmission: A systematic review and meta-analysis. Travel medicine and infectious disease, 36:101751, 2020.

[23] Tao Liu, Jianxiong Hu, Min Kang, Lifeng Lin, Haojie Zhong, Jianpeng Xiao, Guanhao He, Tie Song, Qiong Huang, Zuhua Rong, et al. Transmission dynamics of 2019 novel coronavirus (2019-ncov). 2020a.

[24] Ying Liu, Albert A Gayle, Annelies Wilder-Smith, and Joacim Rocklöv. The reproductive number of covid-19 is higher compared to sars coronavirus. Journal of travel medicine, 2020b.

[25] Bosco Hon-Ming Ma, Terry Cheuk-Fung Yip, Grace Chung-Yan Lui, Mandy Sze-Man Lai, Elsie Hui, Vincent Wai-Sun Wong, Yee-Kit Tse, Henry Lik-Yuen Chan, David Shu-Cheong Hui, Timothy Chi-Yui Kwok, et al. Clinical outcomes following treatment for covid-19 with nirmatrelvir/ritonavir and molnupiravir among patients living in nursing homes. JAMA network open, 6(4):e2310887–e2310887, 2023.

[26] Hemalkumar B Mehta, Shuang Li, and James S Goodwin. Risk factors associated with sars-cov2 infections, hospitalization, and mortality among us nursing home residents. JAMA network open, 4(3):e216315–e216315, 2021.

[27] S Millership and A Cummins. Oseltamivir in influenza outbreaks in care homes: challenges and benefits of use in the real world. Journal of Hospital Infection, 90(4):299–303, 2015.

[28] Arnold S Monto, Klaus Kuhlbusch, Corrado Bernasconi, Bin Cao, Herman Avner Cohen, Emily Graham, Aeron C Hurt, Laurie Katugampola, Takashi Kamezawa, Adam S Lauring, et al. Efficacy of baloxavir treatment in preventing transmission of influenza. New England Journal of Medicine, 392(16):1582–1593, 2025.

[29] Sinead E Morris, Casey M Zipfel, Komal Peer, Zachary J Madewell, Stephan Brenner, Shikha Garg, Prabasaj Paul, Rachel B Slayton, and Matthew Biggerstaff. Modeling the impacts of antiviral prophylaxis strategies in mitigating seasonal influenza outbreaks in nursing homes. Clinical Infectious Diseases, 78(5):1336–1344, 2024.

[30] JY Noh, JY Song, SY Hwang, WS Choi, JY Heo, HJ Cheong, and WJ Kim. Viral load dynamics in adult patients with a (h1n1) pdm09 influenza. Epidemiology & Infection, 142(4):753–758, 2014.

[31] United States Government Accountability Office. Covid-19 in nursing homes: Outbreak duration averaged 4 weeks and was strongly associated with community spread, 2022. URL https://www.gao.gov/assets/820/814143.pdf.

[32] World Health Organization. Coronavirus disease (covid-19): Ventilation and air conditioning in public spaces and buildings. 2021. Accessed: 2025-09-01.

[33] Richard Osei-Yeboah, Stephen Amankwah, Elizabeth Begier, Miranda Adedze, Franklin Nyanzu, Pious Appiah, Jochebed Ode Boakye Ansah, Harry Campbell, Reiko Sato, Luis Jodar, et al. Burden of respiratory syncytial virus (rsv) infection among adults in nursing and care homes: A systematic review. Influenza and other respiratory viruses, 18(9):e70008, 2024.

[34] Zi-Yang Peng, Yun-Ting Hua, Wan-Ting Huang, Jin-Shang Wu, and Huang-Tz Ou. Reduced risks of influenza-associated hospitalization and complications following vaccination among over 2 million older individuals: a nationwide study using target trial emulation framework. BMC medicine, 23(1):157, 2025.

[35] Tin Phan, Ruy M Ribeiro, Gregory E Edelstein, Julie Boucau, Rockib Uddin, Caitlin Marino, May Y Liew, Mamadou Barry, Manish C Choudhary, Dessie Tien, et al. Modeling suggests sars-cov-2 rebound after nirmatrelvir-ritonavir treatment is driven by target cell preservation coupled with incomplete viral clearance. Journal of Virology, 99(3):e01623–24, 2025.

[36] Aurora Pop-Vicas, Momotazur Rahman, Pedro L Gozalo, Stefan Gravenstein, and Vincent Mor. Estimating the effect of influenza vaccination on nursing home residents’ morbidity and mortality. Journal of the American Geriatrics Society, 63(9):1798–1804, 2015.

[37] Lucy Reeve, Elise Tessier, Amy Trindall, Nurin Iwani Binti Abdul Aziz, Nick Andrews, Matthias Futschik, Jessica Rayner, Alexis Didier’Serre, Rebecca Hams, Natalie Groves, et al. High attack rate in a large care home outbreak of sars-cov-2 ba. 2.86, east of england, august 2023. Eurosurveillance, 28(39):2300489, 2023.

[38] Jeanine JS Rutten, Anouk M van Loon, Janine van Kooten, Laura W van Buul, Karlijn J Joling, Martin Smalbrugge, and Cees MPM Hertogh. Clinical suspicion of covid-19 in nursing home residents: symptoms and mortality risk factors. Journal of the American Medical Directors Association, 21(12):1791–1797, 2020.

[39] Isabel San Martín-Erice, Paula Escalada-Hernández, Cristina García-Vivar, Sara Furtado-Eraso, Leticia San Martín-Rodríguez, and Nelia Soto-Ruiz. How did covid-19 lockdown impact the health of older adults in nursing homes? a scoping review. BMC geriatrics, 24(1):760, 2024.

[40] Joyce Simard and Ladislav Volicer. Loneliness and isolation in long-term care and the covid-19 pandemic. Journal of the American Medical Directors Association, 21(7):966–967, 2020.

[41] GJB Sousa, TS Garces, VRF Cestari, R. Florêncio, TMM Moreira, and MLD Pereira. Mortality and survival of covid-19. Epidemiology & Infection, 148:e123, 2020.

[42] Momoe Utsumi, Kiyoko Makimoto, Nahid Quroshi, and Nobuyuki Ashida. Types of infectious outbreaks and their impact in elderly care facilities: a review of the literature. Age and ageing, 39(3):299–305, 2010.

[43] Carline Van den Dool, Eelko Hak, Marc JM Bonten, and Jacco Wallinga. A model-based assessment of oseltamivir prophylaxis strategies to prevent influenza in nursing homes. Emerging Infectious Diseases, 15(10):1547, 2009.

[44] Marianne AB van der Sande, Adam Meijer, Fatmagül Şen-Kerpiclik, Remko Enserink, Herman JM Cools, Piet Overduin, José M Ferreira, Marie-José Veldman-Ariessen, and Peppie study group. Effectiveness of post-exposition prophylaxis with oseltamivir in nursing homes: a randomised controlled trial over four seasons. Emerging Themes in Epidemiology, 11(1):13, 2014.

[45] Catharina E van Ewijk, Elizabeth I Hazelhorst, Susan JM Hahné, and Mirjam J Knol. Covid-19 outbreak in an elderly care home: very low vaccine effectiveness and late impact of booster vaccination campaign. Vaccine, 40(46):6664–6669, 2022.

[46] Philippe Vanhems, Alain Barrat, Ciro Cattuto, Jean-François Pinton, Nagham Khanafer, Corinne Régis, Byeul-a Kim, Brigitte Comte, and Nicolas Voirin. Estimating potential infection transmission routes in hospital wards using wearable proximity sensors. PloS one, 8(9): e73970, 2013.

[47] Alexandre Worcel, Bougacha M Ali, Sonia Ramos-Pascual, Patrick Stirling, and Francois G Chary. Low mortality from covid-19 at a nursing facility in france following a combined preventive and active treatment protocol. Annals of Palliative Medicine, 10(11):112881300– 112811300, 2021.

[48] Yu Wu, Liangyu Kang, Zirui Guo, Jue Liu, Min Liu, and Wannian Liang. Incubation period of covid-19 caused by unique sars-cov-2 strains: a systematic review and meta-analysis. JAMA network open, 5(8):e2228008–e2228008, 2022.

[49] JLY Yip, Smita Kapadia, A Ahmed, and S Millership. Outbreaks of influenza-like illness in care homes in the east of england: impact of variations in neuraminidase inhibitor provision. Public Health, 162:98–103, 2018.

[50] Hind Zaaraoui, Clarisse Schumer, Xavier Duval, Bruno Hoen, Lulla Opatowski, and Jérémie Guedj. Modelling the effectiveness of antiviral treatment strategies to prevent household transmission of acute respiratory viruses. PLOS Computational Biology, 20(12):e1012573, 2024.

[51] Chenliang Zhou, Tianfang Zhang, Haotang Ren, Shanshan Sun, Xia Yu, Jifang Sheng, Yu Shi, and Hong Zhao. Impact of age on duration of viral rna shedding in patients with covid-19. Aging (Albany NY), 12(22):22399, 2020.

