## Supplementary Methods, Tables and Figure for "Bridging individual and population dynamics of acute respiratory viruses to optimize antiviral interventions in nursing homes"

November 14, 2025

#### Methods

##### Models of viral load dynamics

###### Covid-19

The model used for SARs-CoV-2 is described below. Modeling and calibration is described in Zaaraoui et al. [2024], parameters are presented in Table 1:

$$\frac{dT}{dt} = -\beta TV - \phi TF + \rho R$$
$$\frac{dR}{dt} = \phi FT - \rho R$$
$$\frac{dI_1}{dt} = \beta VT - kI_1$$
$$\frac{dI_2}{dt} = kI_1 - \Delta(t)I_2$$
$$\frac{dV_I}{dt} = \pi\mu(1 - \frac{F}{F+\theta})I_2 - cV_I$$
$$\frac{dV_{NI}}{dt} = \pi(1-\mu)(1 - \frac{F}{F+\theta})I_2 - cV_{NI}$$
$$\frac{dF}{dt} = I_2 - d_f F$$
$$\text{where } \begin{cases} \Delta(t) = \delta_1 & t < \tau \\ \Delta(t) = \delta_2 \times (1 - 0.75 \times 1_{age>65}) & t \geq \tau. \end{cases}$$

where T, R etc

###### Influenza

Mathematical model calibration for influenza A virus (IAV) infection was performed using individual viral load trajectories extracted from the dataset published in Asher et al. [2023]. This dataset compiles viral kinetic data from the placebo arms of three phase II/III clinical trials evaluating antiviral efficacy, including Ison et al. [2020] and Hayden et al. [2018]. Only placebo recipients were retained in our analysis to characterize the natural course of within-host viral replication and clearance without pharmacologic interference. For each subject, nasopharyngeal viral loads were log-transformed and aligned by time since symptom onset. The resulting dataset provided high-resolution time series of viral kinetics, which were used to estimate the parameters (see Table 1) governing viral replication rate, infected cell death, and immune-mediated clearance through nonlinear modeling. Model fitting was performed using maximum likelihood estimation, assuming log-normal residuals, and parameter uncertainty was quantified through bootstrap resampling. The model is described below:

$$
\begin{aligned}
\frac{dT}{dt} &= -\beta T V \\
\frac{dI_1}{dt} &= \beta T V - k I_1 \\
\frac{dI_2}{dt} &= k I_1 - \Delta(t) I_2 \\
\frac{dV}{dt} &= \pi I_2 - c V \\
&\text{where } \begin{cases} \Delta(t) = \delta_1 & t < \tau \\ \Delta(t) = \delta_2 \times (1 - 0.75 \times 1_{age>65}) & t \geq \tau. \end{cases}
\end{aligned}
$$

where T, R etc

##### RSV

RSV model's parameters estimation (see Table 1) was based on experimental human challenge data obtained through a collaboration with hVivo. The dataset included 369 healthy adults aged 18–55 years who participated in 11 controlled challenge studies conducted between 2013 and 2020. All participants were pre-screened for low RSV-neutralizing antibody titers and were inoculated intranasally with approximately $4 \log_{10}$ PFU/mL of the RSV Memphis-37b strain. Beginning on day 2 post-inoculation, 36–48 hours after exposure, both infectious virus and total viral RNA were measured twice daily until viral clearance. Only placebo or untreated individuals were retained for analysis to capture the natural viral kinetics. The RSV within-host dynamics were described by the following model:

$$
\begin{aligned}
\frac{dT}{dt} &= -\beta T V - \phi T F + \rho R \\
\frac{dR}{dt} &= \phi F T - \rho R \\
\frac{dI_1}{dt} &= \beta V T - k I_1 \\
\frac{dI_2}{dt} &= k I_1 - \Delta(t) I_2 \\
\frac{dV}{dt} &= \pi (1 - \frac{F}{F + \theta}) I_2 - c V \\
\frac{dF}{dt} &= I_2 - d_f F \\
&\text{where } \begin{cases} \Delta(t) = \delta_1 & t < \tau \\ \Delta(t) = \delta_2 \times (1 - 0.75 \times 1_{age>65}) & t \geq \tau. \end{cases}
\end{aligned}
$$

Parameters were estimated by computing the maximum likelihood estimator using the stochastic approximation expectation-maximization (SAEM) algorithm implement in Monolix 2023R1.

| Parameters | Description | SARS-CoV-2 | IAV<br>fixed effect(random effect)* | RSV |
|---|---|---|---|---|
| $\beta$ | Infection rate | $1.89 \times 10^{-5}$ (0.699) | $1.12 \times 10^{-5}$ (1.12) | $1.02 \times 10^{-6}$ (0.615) |
| $\tau$ | Adaptive response time | 14.47 (0.401) | 3.63 (0.149) | 7.28 (0.116) |
| $\delta_1$ ($d^{-1}$) | Loss rate of infected cells at $t < \tau$ | 0.976 (0.231) | 0.464 (0.35) | 0.103 (0.291) |
| $\delta_2$ ($d^{-1}$) | Loss rate of infected cells at $t \geq \tau$ | 3.56 (0.27) | 7.91 (0.823) | 6.57 (0.999) |
| $\pi$ ($cp \times mL^{-1}d^{-1}$) | Rate of viral production | $6.43 \times 10^5$ ( - ) | 34 ( - ) | $2.29 \times 10^4$ ( - ) |
| $c$ ($d^{-1}$) | Virion clearance rate | 10 ( - ) | 10 ( - ) | 10 ( - ) |
| $k$ ($d^{-1}$) | Rate of transition to productively infected cells | 4 ( - ) | 8 ( - ) | 4 ( - ) |
| $\mu$ | Proportion of infectious virus | 0.0001 ( - ) | NA | NA |
| $\phi$ ($d^{-1}$) | Refractory rate of target cells | 0.00124 (0.266) | NA | $2.79 \times 10^{-5}$ (5.84) |
| $d_F$ ($d^{-1}$) | Loss rate of effectors | 3.03 (0.83) | NA | 2 ( - ) |
| $\theta$ | Half-maximal concentration of effectors | 1720 (1.55) | NA | 9140 (1.26) |
| $\rho$ ($d^{-1}$) | Rate of transition from refractory to susceptible cells | 0.00431(0.123) | NA | 0.34 ( - ) |
| $\epsilon$ | Antiviral treatment efficacy | 0.94 ( - ) | 0.92 ( - ) | 0.84 ( - ) |
| **Risk of hospitalization** | **Description** | | | |
| $\nu$ | Shape parameter | 0.8538 | 3.2333 | 8.35 |
| $\lambda$ ($cp/mL$) | Viral dynamic effect | $5.26 \times 10^{-4}$ | 1.1139 | 1.943 |
| $\frac{1}{\gamma}$ | Scale parameter | $5 \times 10^{-4}$ | 0.00451 | 0.0089 |
| **Virus transmission** | **Description** | | | |
| $m$ (mL/copies) | $p(t) = 1 - e^{m \times V(t)^{0.49}}$ | $m = 16 \times 10^{-7}$ | $m = 21 \times 10^{-7}$ | $m = 33.7 \times 10^{-7}$ |

Table 1: **List of model parameters. Parameter definition and values for the models of viral dynamic, transmission probability and risk of severe disease profiles***. **All random parameters are assumed to have a log-normal distribution. Parameters with no random effects (same parameter value in all individuals) are indicated with "-". cp = copies.**

#### Modelling the impact of antiviral treatment on viral load

The antiviral treatments for the three viruses are assumed to reduce viral production with an efficacy rate denoted by $\epsilon$. The treatment, initiated at time $t_x$, is incorporated into the viral dynamics model by modifying the equations as follows:

- For SARS-CoV-2 model:

$$\frac{dV_I}{dt} = (1 - \epsilon \times 1_{t \geq t_x})\pi\mu(1 - \frac{F}{F+\theta})I_2 - cV_I$$
$$\frac{dV_{NI}}{dt} = (1 - \epsilon \times 1_{t \geq t_x})\pi(1-\mu)(1 - \frac{F}{F+\theta})I_2 - cV_{NI}$$

- For influenza model:

$$\frac{dV}{dt} = (1 - \epsilon \times 1_{t \geq t_x})\pi I_2 - cV$$

- For RSV model:

$$\frac{dV}{dt} = (1 - \epsilon \times 1_{t \geq t_x})\pi(1 - \frac{F}{F+\theta})I_2 - cV$$

The value of this efficacy parameter depends on the relative reduction in the risk of severe disease for each virus. For a 50% reduction in severe disease, the corresponding $\epsilon$ values are reported in Table 1. Reductions of 80% and 30% correspond to $\epsilon$ values of 0.99 and 0.7 respectively for SARS-CoV-2, 0.9999, with a treatment initiation time at symptom time onset, and 0.9 for IAV respectively, and 0.99 and 0.62 for RSV, respectively.

#### Simulations of outbreaks in nursing home

##### Settings of the simulation study

For each scenario, simulations were run by fixing the following conditions and hypotheses:

- 1000 nursing homes
- Nursing homes have size $N = 120 = N_r + N_s$, with $N_r = 68$ the number of residents and $N_s$ is the number of NHW, corresponding to a ratio of 0.75 staff/resident. This figure is representative of the average size of nursing homes in France DREES [2023]
- Time step $t$ is 1 day.
- Initialization of the model is set at $t_0 = 1$ assumed to be the infection time of one index $i_0$. All other individuals are assumed to be susceptible in the nursing home,
- The minimal duration of contact leading to transmission is 5 minutes,
- Transmission risk during contact saturates at 30 minutes,
- The simulation lasts 15 weeks.

##### Contact matrix

Interactions between individuals within nursing homes are defined through two empirical contact matrices parameterized based on a contact study Duval et al. [2018] conducted in a Long Term Care Facility (LTCF) in France:

- $F$, the average group-level frequency matrix. $F$ is a $2 \times 2$ matrix, each entry representing the average number of distinct individuals contacted per day per category. We set: 7.1 residents-residents contacts, 3.9 residents-NHW contacts, 8 NHW-residents contacts and 4.9 NHW-NHW contacts,

- $D$, the group-level matrix of mean cumulative daily contact durations. $D$ is of size $2 \times 2$. It defines the daily cumulated duration of contacts between all pairs of individuals from different groups: residents-residents: 31.7 minutes/day; residents-NHW: 10.3 min; NHW-residents: 13.6 min; and NHW-NHW: 18.6 min.

In a nursing home of size $N = N_r + N_s$, with $N_r$ residents and $N_s$ staff (NHW), we stochastically generated, for each day $t$, the daily individual-level contacts $N \times N$ stochastic matrices:

- $\mathcal{F}_t$, the randomized contact frequency matrix: each element $\mathcal{F}_{(i,j),t} \in \{0, 1\}$ indicates the presence (1) or absence (0) of contact between individuals $i$ and $j$.This matrix is re-computed each day $t$, and $\mathcal{F}_{(i,j),t}$ is initialized (day $t = 1$) from a Bernoulli distribution. $\mathcal{B}$ with parameter $F_{g(i),g(j)} \times \frac{1}{N_{g(j)}}$, where $g(j) = r$ if $j$ is a resident, $g(j) = s$ if $j$ is a NHW, and $N_{g(j)}$ represents the size of the $g(j)$'s group:

$$\mathcal{F}_{(i,j),t} \sim \mathcal{B}\left(F_{g(i),g(j)} \times \frac{1}{N_{g(j)}}\right), \tag{1}$$

- $\mathcal{D}_t$, the randomized cumulative contact duration matrix: each element $\mathcal{D}_{(i,j),t}$ is Normal-randomly sampled on each day with mean $D_{g(i),g(j)}$, assuming a standard deviation of 3 days:

$$\mathcal{D}_{(i,j),t} \sim \mathcal{N}\left(D_{g(i),g(j)}, 9\right), \tag{2}$$

From these quantities, we compute, for each day $t$, the effective time-in-contact matrix daily $C(t)$. Each element denotes the cumulative contact duration between individuals $i$ and $j$, computed by:

$$C_{i,j}(t) = \mathcal{F}_{(i,j),t} \times \mathcal{D}_{(i,j),t}, \tag{3}$$

$C_{i,j}(t) = 0$ if no contact between the potential transmitter $i$ and the potential receiver $j$ occurred at time step $t$.

##### Recurrent contacts

To capture the recurrent contacts effect, the previous matrix was adjusted. We define the effective contact probability $\omega$, between individuals $i$ and $j$, which incorporates the past contact behavior between individuals, thereby replacing the uniform distribution $\frac{1}{N_{g(j)}}$ with a time-dependent weight $\omega_{i,j}(t)$ in the $\mathcal{F}$ matrix (see (6)). The evolution of $\omega(t)$ over time provides information on dynamics of evolution groups of contacts from individuals' admission in the nursing home, and is defined by:

$$\omega_{i,j}(t) = \frac{\overline{n}_{i,j}(t)}{\sum_{k=1}^{S_j} \overline{n}_{i,k}(t)} \tag{4}$$

where $\overline{n}_{i,j}(t)$ is the exponential average of past contact events Brown [2004] between $i$ and $j$, denoted by $ct_{i,j}$, from the start of the study, assigning progressively lower weights to older interactions. When $t > 0$, this quantity is recursively defined to reflect recent contact patterns more strongly by:

$$\begin{aligned}\overline{n}_{i,j}(t) &= \alpha \times ct_{i,j}(t-1) + (1-\alpha)\overline{n}_{i,j}(t-1) \qquad (5)\\ &= \alpha \times ct_{i,j}(t-1) + \alpha \sum_{d=1}^{t-2} (1-\alpha)^{t-1-d} \overline{n}_{i,j}(d)\end{aligned}$$

$\sum_{k=1}^{N_j} \overline{n}_{i,k}(d)$ is the exponential mean number of contacts between $i$ and all $k$ individuals of the $j's$ category of size $N_j$ until day $t$. The parameter $\alpha \in [0, 1]$ controls the recurrence of contacts.

When $\alpha = 0$, contacts are uniformly distributed across all individuals in the nursing home, indicating no memory of previous interactions. In contrast, as $\alpha \to 1$, the probability of repeated contact with the same individual on consecutive days approaches 1, reflecting highly recurrent contact patterns. Although the contact network may not strictly reach a steady state, we fixed the contact pattern to that observed after six months of simulated interactions. Empirical evidence further indicates that over 85% of resident or NHW contacts are recurrent Duval [2019], Vanhems et al. [2013], corresponding to $\alpha = 0.11$ in (5). This contact configuration, denoted by $\omega$, is then incorporated into $\mathcal{F}$ as follows:

$$\mathcal{F}_{i,j} \sim \mathcal{B}\left(F_{g(i),g(j)} \times \omega_{i,j}\right), \tag{6}$$

**Transmission matrix simulation**

The transmission risk is modeled through the following three matrices of size $N \times N$:

- $I(t)$, the infection vector: it defines the infection status (of value 0 or 1) of the individuals in row $i \in [|1, N|]$ at time step $t$,
- $S(t)$, the susceptibility matrix: it defines the susceptibility status of the individuals (of value 0 or 1) in row $i \in [|1, N|]$ at time step $t$. An individual is susceptible if he is not infected yet since the beginning of the outbreak,
- $P(t)$, the effective transmission probability matrix at time step $t$: each of its element is computed by the daily cumulative transmission defined by $P_{i,j}(t)$ in Eq.(4) of the main paper. If the contact lasts less than 5 minutes, the probability of transmission is fixed to 0. In our model, contact durations are defined discretely by intervals of 5 minutes.

Status vectors are defined as follows. On the one hand, we have the infection vector $I(t)$ that defines the infection status of all individuals in row $i$ at time step $t$:

- $I_i(t) = 1,\ \forall j \in [|1, N|]$ if $i$ has been infected before or at day $t$
- $I_i(t) = 0,\ \forall j \in [|1, N|]$ if $i$ is not infected until day $t$

On the other hand, $S(t)$ defines the susceptibility vector of the individuals $j$ since the beginning of the current outbreak:

- $S_j(t) = 1,\ \forall j \in [|1, N|]$ if $j$ is not yet infected up to time $t$
- $S_j(t) = 0,\ \forall j \in [|1, N|]$ if $j$ has been already infected before or at time $t$

The transmission within the nursing home is modeled through the matrix model that computes the infection probability at each time step $t$, $T(t)$ (7). In this matrix, all $N$ individuals present in the nursing home are represented by rows, where each individual may be potentially infectious and transmit the pathogen. The first $N_r$ rows ($i = 1, \ldots, N_r$, with $N_r = 68$ in our case) correspond to residents, while the subsequent $N_s$ rows ($i = N_r + 1, \ldots, N$) correspond to nursing home staff (NHW). Similarly the $N$ columns represent all nursing home's individuals as potential susceptible. The first $N_r$ columns representing residents and the following $N_s$ columns the staff. A given $(i, j)$ cell of the matrix informs about the transmission of individual $j$ by individual $i$.

Therefore the simulated daily risk of transmission $T$ from individual $i$ to individual $j$ at time $t$ is defined by the matrix $T$:

$$T_{i,j}(t) = \left(1 - (1 - p_i(t))^{\left\lfloor \frac{\min\left(C_{i,j}(t),30\right)}{5} \right\rfloor}\right) \times I_i(t) \times S_j(t) \tag{7}$$

The infection of an individual $j$ (from the $j^{th}$ column) by the infected individual $i$ is not automatic. It depends on the contact matrix, and therefore on the probability of the transmission $P$ where $P_{i,j}(t) = 1 - (1 - p_i(t))^{\min\left(\left\lfloor \frac{C_{i,j}(t)}{5} \right\rfloor, \left\lfloor \frac{30}{5} \right\rfloor\right)}$. Note that:

$$P_{i,j}(t) = \begin{cases} 0, & \text{if } j = i, \\ 0, & \text{if no contact exists between } i \text{ and } j \text{ at time } t, \\ 0, & \text{if } i \text{ is not infected or recovered.} \end{cases}$$

The updated infection status of an individual $j$ at time $t+1$ depends on all the potential transmission from any individual $i$ in the nursing home:

$$I_j(t+1) \sim \max_{i \in [|1,N|],\ i \neq j} \mathcal{B}(T_{i,j}) \tag{8}$$

where $\mathcal{B}(T_{i,j})$ is the Bernoulli random variable of parameter $T_{i,j}$. Note that if $T_{i,j} = 0$, therefore $I_j(t+1) = 0$ i.e., the individual $j$ is not infected at time $t+1$.

#### Individuals' infection time

A time of infection vector $\tau$ is also computed at each time $t$. If an individual $j$ is infected at time $t$, the vector $\tau$ is updated as following:

$$\tau_j(t) = \begin{cases} t, & \text{if individual } j \text{ is newly infected at time } t, \\ \tau_j(t-1), & \text{if individual } j \text{ was already infected before } t, \\ 7 \times 15 + 1, & \text{if individual } j \text{ is still susceptible.} \end{cases}$$

which also can be rewritten by:

$$\tau_j(t+1) = t \times I_j(t+1) + (1 - I_j(t+1)) \times \tau_j(t) \tag{9}$$

This vector records the infection times of all individuals. At time $t$, it assigns the current infection time to newly infected individuals, while retaining the previously assigned infection times for those already infected before $t$. For individuals who remain susceptible throughout the simulation, the infection time is set to $\tau_j = 7 \times 15 + 1$, corresponding to a censoring value beyond the simulation horizon of 15 weeks after the index infection.

| **Virus and antiviral treatment** | | | |
|---|---|---|---|
| | **SARS-CoV-2** | **IAV** | **RSV** |
| Time to peak viral load (days) | 4<br>(3 - 6) | 2<br>(1 - 3) | 5<br>(2 - 8) |
| Proportion of virus shed during the pre-symptomatic phase | 17% | 75% | 29% |
| Proportion of asymptomatic infection | 50% | 50% | 50% |
| Duration of the incubation period (days) | 4<br>(3 - 5) | 2<br>(1 - 3) | 5<br>(2 - 8) |
| Risk of severe disease in symptomatic individuals | 25% | 12% | 12% |
| Efficacy of treatment against severe disease<br>(after symptom onset) | 50% | 50% | 50% |

| **Contact and transmission** | | | |
|---|---|---|---|
| | **Resident→Resident** | **Resident $\leftrightarrows$ NHW** | **NHW→NHW** |
| Groups size ($N_r$ and $N_s$) | | $N_r = 68 \leftrightarrows N_s = 52$ | |
| Number of recurrent contacts (over 10 days) | 3.2 | 2.1 | 2.3 |
| Proportion of contacts >30 minutes | $> 80\%$ | $< 1\%$ | 45% |
| Basic reproduction number ($R0$) | 3 | 1.5 | 1.5 |

| **Interventions** | | |
|---|---|---|
| | **Targeted individuals** | **Duration** |
| Mask facing of all NHW[(1)] | At outbreak detection | Until end of the outbreak |
| Mask facing of symptomatic NHW[(1)] | 0-5 days after symptom onset | Until recovery |
| Isolation[(2)] of symptomatic residents | 0-5 days after symptom onset | Until recovery |
| Isolation of contact residents | at detection of the index case | 10 days |
| Isolation of all residents | All residents, 0-5 days after any second symptomatic detection | Until end of the outbreak |
| Treatment of symptomatic residents | 0-5 days after symptom onset | Until recovery |
| Treatment of contact infected residents | at detection of the index case | Until recovery |
| Treatment of all contact residents | at detection of the index case | Pep short: 5 days<br>Pep long: Until end of the outbreak |

| **Model outcomes** | | | |
|---|---|---|---|
| | **SARS-CoV-2** | **IAV** | **RSV** |
| Efficacy of treatment against severe disease<br>(after symptom onset) | 50% | 50% | 50% |
| Efficacy of treatment against severe disease<br>(before symptom onset) | 85% | 75% | 73% |
| Efficacy of treatment against severe disease (PEP) | 99% | 90% | 84% |
| Efficacy of treatment against transmission<br>(after symptom onset) | 20% | 30% | 20% |
| Efficacy of treatment against transmission<br>(before symptom onset) | 80% | 67% | 75% |
| Efficacy of treatment against transmission (PEP) | 98% | 96% | 85% |

Table 2: Main assumptions and outcomes of the model. [(1)] Mask facing reduces transmission/infection by 80%. [(2)] Isolation reduces contacts with residents and NHW by 100 and 80%.

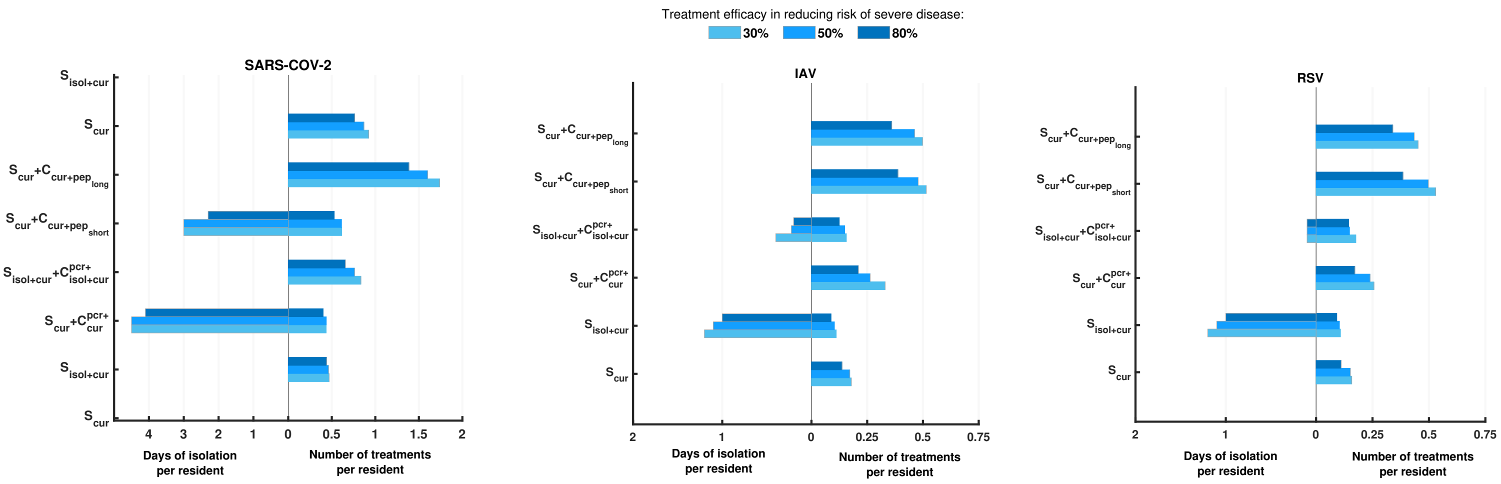


Figure 1: **Mean number of isolation days and number of treatments per resident for different treatment efficacy.**

### References

Jason Asher, Annabelle Lemenuel-Diot, Matthew Clay, David P Durham, Luis Mier-y Teran-Romero, Carlos J Arguello, Sebastien Jolivet, Diana Y Wong, Klaus Kuhlbusch, Barry Clinch, et al. Novel modelling approaches to predict the role of antivirals in reducing influenza transmission. *PLoS Computational Biology*, 19(1):e1010797, 2023.

Robert Goodell Brown. *Smoothing, forecasting and prediction of discrete time series.* Courier Corporation, 2004.

DREES. L'enquête auprès des établissements d'hébergement pour personnes âgées (ehpa). `https://data.drees.solidarites-sante.gouv.fr/explore/dataset/587_l-enquete-aupres-des-etablissements-d-hebergement-pour-personnes-agees-ehpa/information/`, 2023.

Audrey Duval. *Comprendre et contrôler la transmission des bactéries multirésistantes par l'analyse et la modélisation des réseaux d'interactions interindividuelles en milieu hospitalier.* PhD thesis, Université Paris Saclay (COmUE), 2019.

Audrey Duval, Thomas Obadia, Lucie Martinet, Pierre-Yves Boëlle, Eric Fleury, Didier Guillemot, Lulla Opatowski, and Laura Temime. Measuring dynamic social contacts in a rehabilitation hospital: effect of wards, patient and staff characteristics. *Scientific reports*, 8(1):1686, 2018.

Frederick G Hayden, Norio Sugaya, Nobuo Hirotsu, Nelson Lee, Menno D de Jong, Aeron C Hurt, Tadashi Ishida, Hisakuni Sekino, Kota Yamada, Simon Portsmouth, et al. Baloxavir marboxil for uncomplicated influenza in adults and adolescents. *New England Journal of Medicine*, 379(10):913–923, 2018.

Michael G Ison, Simon Portsmouth, Yuki Yoshida, Takao Shishido, Melissa Mitchener, Kenji Tsuchiya, Takeki Uehara, and Frederick G Hayden. Early treatment with baloxavir marboxil in high-risk adolescent and adult outpatients with uncomplicated influenza (capstone-2): a randomised, placebo-controlled, phase 3 trial. *The Lancet Infectious Diseases*, 20(10):1204–1214, 2020.

Philippe Vanhems, Alain Barrat, Ciro Cattuto, Jean-François Pinton, Nagham Khanafer, Corinne Régis, Byeul-a Kim, Brigitte Comte, and Nicolas Voirin. Estimating potential infection transmission routes in hospital wards using wearable proximity sensors. *PloS one*, 8(9): e73970, 2013.

Hind Zaaraoui, Clarisse Schumer, Xavier Duval, Bruno Hoen, Lulla Opatowski, and Jérémie Guedj. Modelling the effectiveness of antiviral treatment strategies to prevent household transmission of acute respiratory viruses. *PLOS Computational Biology*, 20(12):e1012573, 2024.
